# NEWS2 and laboratory predictors correlated with clinical deterioration in hospitalised patients with COVID-19

**DOI:** 10.1101/2021.01.17.21249878

**Authors:** Gulsah Tuncer, Serkan Surme, Osman Faruk Bayramlar, Hatice Kubra Karanalbant, Betul Copur, Meltem Yazla, Esra Zerdali, Inci Yilmaz Nakir, Ayse Kurt Cinar, Ahmet Buyukyazgan, Hatice Balli, Yesim Kurekci, Serap Simsek Yavuz, Mehmet Mesut Sonmez, Gonul Sengoz, Filiz Pehlivanoglu

**Affiliations:** Department of Infectious Diseases and Clinical Microbiology, Haseki Training and Research Hospital, Istanbul, Turkey; Department of Public Health, Bakirkoy District Health Directorate, Istanbul, Turkey; Department of Infectious Diseases and Clinical Microbiology, Istanbul Faculty of Medicine, Istanbul University, Istanbul, Turkey; Department of Orthopaedic Surgery and Traumatology, Haseki Training and Research Hospital, Istanbul, Turkey

**Keywords:** COVID-19, NEWS2, in-hospital mortality, procalcitonin, albumin, neutrophil/lymphocyte ratio

## Abstract

**Background:** We aimed to determine prognostic values of NEWS2 and laboratory parameters during the first week of COVID-19.

**Methods:** All adult patients who were hospitalized for a confirmed COVID-19 between the 11th of March and the 11th of May 2020 were retrospectively included. To evaluate the factors in prognosis which are admission to intensive care unit (ICU) and in-hospital death, univariate logistic regression analysis was performed at admission (D0), at day-3 (D3), day-5 (D5), and day-7 (D7). Additionally, receiver operating characteristic (ROC) analyses were performed.

**Results:** Overall, 611 patients were included. Clinical deterioration was observed in 79 (12.9%) patients during hospitalisation, 36 (5.9%) during the first three days, 54 (8.8%) during the first five days, and 62 (10.1%) during the first week of hospitalisation. Our results showed that NEWS2, procalcitonin, neutrophil/lymphocyte ratio (NLR), and albumin were the best predictors for clinical deterioration at D0, D3, D5, and D7. Procalcitonin had the highest odds ratio for clinical deterioration on all days in univariate analysis. ROC analyses showed that NEWS2 at D7, procalcitonin at D5, albumin at D7, and NLR at D5 had highest AUC values. Additionally, we detected a strong correlation between NEWS2 and laboratory parameters including neutrophil, lymphocyte, NLR, platelet/lymphocyte ratio, CRP, procalcitonin, ferritin, and urea on all days.

**Conclusion:** This study provides a list of several laboratory parameters correlated with NEWS2 and potential predictors for ICU admission or in-hospital death during the clinical course of COVID-19. Dynamic monitoring of NEWS2 and laboratory parameters is vital for improving clinical outcomes.

## 1. INTRODUCTION

The COVID-19 pandemic due to the SARS-CoV-2 virus causes high rates of mortality, morbidity, longer duration of hospitalization and increased need for intensive care unit (ICU) admission (1). Improving critical care patient flow is crucial for high quality care to severe cases. Therefore, we need to predict clinical deterioration in patients with COVID-19, in order to hospitalize the patients and admit to ICU, when necessary. The National Institute for Health and Care Excellence recommend The National Early Warning Score 2 (NEWS2) to predict the risk for clinical deterioration in patients with COVID-19 (2, 3). NEWS2 is a simple scoring system including physiological parameters and vital signs (respiratory rate, oxygen saturation, systolic blood pressure, heart rate, level of consciousness, temperature and supplemental oxygen dependency) used to predict the risk for acute deterioration including sepsis (4, 5). An increasing number of studies have assessed the parameters in NEWS2 score for severe COVID-19 illness (6). However there is a lack of knowledge about the predictive value of NEWS2, despite some studies focus on NEWS2 and related scores (3).

In this study, we aimed to determine prognostic value of NEWS2 and laboratory parameters in COVID-19 patients. Additionally, the correlation between NEWS2 and laboratory parameters at admission, D3, D5, and D7 during the clinical course of COVID-19 were evaluated.

## 2. PATIENTS AND METHODS

### 2.1. Study Design and Patients

In this retrospective and single-center study, all adult patients (≥18 years old) who were hospitalized for a laboratory confirmed COVID-19 between the 9th of March and the 8th of May 2020 were included. SARS-CoV-2 testing was performed by real-time reverse transcription-polymerase chain reaction (RT-PCR) of samples collected by nasopharyngeal and/or oropharyngeal swabs.

Patients with COVID-19 requiring hospitalisation were included in the study. Outpatients and asymptomatic patientswere excluded. Also, we excluded patients if oropharyngeal or nasopharyngeal swab samples were repeatedly negative for SARS-CoV-2 by RT-PCR. Our primary outcome was the occurence of clinical deterioration defined as a composite of ICU admission during hospitalisation or in-hospital death.

### 2.2 Data collection

Epidemiological and demographic characteristics, clinical, laboratory, radiological findings, and outcomes were collected from medical records. Vital signs including respiratory rate, peripheral capillary oxygen saturation, heart rate, blood pressure, body temperature, and consciousness (Glasgow Coma Scale) were recorded. Laboratory parameters including albumin, C-reactive protein (CRP), procalcitonin, haemoglobin, hematocrit, neutrophil count, lymphocyte count,, platelet count, neutrophil/lymphocyte ratio (NLR), platelet/lymphocyte ratio (PLR), urea, ferritin, albumin, fibrinogen, d-dimer, aspartate aminotransferase (AST), and alanin aminotransferase (ALT) were included. The NEWS2 score was calculated.

### 2.3 Statistical analysis

Quantitative variables are expressed as mean & standard deviation if they contain continuous and normal distributed data. If the data was not distributed normally, median and interquartile range (IQR) were used. If they contain categorical data, they are expressed as percentage (%) and frequency (n). Comparison of qualitative variables was analyzed with Pearson Chi square test. The normal distribution questioning the necessity of using the parametric test was examined by Kolmogrov-Smirnov, Shapiro Wilk, Curtosis - Skewness Tests and Box Plot Distribution. When normally distributed data could not be determined and non-parametric tests and spearman correlation were used. Kruskal Wallis Test was used for the analysis of continuous and more than two independent non-parametric groups (Bonferroni correction was used when necessary) and Mann Whitney Test was used for post-hoc analysis. To evaluate the factors in prognosis which are admission ICU and in-hospital death, univariate logistic regression analysis was performed. Afterwards, these dependent groups were handled one by one, ROC curves were drawn and cut-off values, sensitive and specificity of cut-off values, and Area Under the Curve (AUC), were demonstrated. To predict clinical deterioration, the prognostic accuracy of NEWS2 and laboratory parameters was evaluated for:

1. NEWS2 and laboratory parameters associated with the clinical deterioration at D0
2. NEWS2 and laboratory parameters associated with the clinical deterioration at D3 after excluding from the analysis patients with clinical deterioration within the first three days of hospitalization.
3. NEWS2 and laboratory parameters associated with the clinical deterioration at D5 after excluding from the analysis patients with clinical deterioration within the first five days of hospitalization.
4. NEWS2 and laboratory parameters associated with the clinical deterioration at D7 after excluding from the analysis patients with clinical deterioration within the first seven days of hospitalization.

The results were evaluated in 95% confidence interval and statistical significance level was defined as p <0.05. The analyzes were performed using IBM SPSS - 21 (Statistical Package for Social Sciences, Chicago, IL, USA)

## 3. RESULTS

### 3.1 General Characteristics

Overall, 611 patients were included. Of whom, 329 (53.8%) were male, the mean age was 52.53±15.07 years. 73 patients (11.9%) were admitted to the ICU. In-hospital death occurred in 46 (7.5%) patients. Clinical deterioration was observed in 79 patients (12.9%) during hospitalisation, 36 (5.9%), during the first three days, 54 (8.8%) during the first five days, and 62 (10.1%) during the first week of hospitalisation. NEWS2 was calculated at admission (D0), D3, D5 and D7 of hospitalization. Patients were stratified into three risk groups: low risk from 0 to 4; medium risk from 5 to 6 and high risk above 7. Of 611 patients, 505 (82.7%) at D0, 411 (91.9%) at D3, 375 (92.2%) at D5, 284 (93.8%) at D7 had a NEWS2 score<7. The median length of hospital stay was 8.9 days and 332 patients (54.3%) who did not have fever and did not need oxygen in the last 48-72 hours and meet the criteria for home monitoring were discharged within the first 7 days. Characteristic of patient, NEWS2 value and NEWS2 class at D0, D3, D5 and D7, are represented in Supp. Table.

**Suppl. Table.** Characteristics, NEWS2 value anda class of patients hospitalized for COVID-19

|  | Overall (n: 611) |
| --- | --- |
| Age, mean ±SD | 52.53±15.07 |
| Sex |  |
| Male, n (%) | 329 (53.8%) |
| **Serious COVID-19 risk factors, n (%)** |  |
| Age >65 years old, n (%) | 131 (21.3%) |
| Diabetes mellitus, n (%) | 170 (27.6%) |
| Hypertension | 192 (31.2%) |
| Cardiovascular diseases, n (%) | 13 (2.1%) |
| Cerebrovascular diseases, n (%) | 4 (0.6%) |
| Chronic kidney failure, n (%) | 34 (5.5%) |
| Malignancy, n (%) | 10 (1.2%) |
| Chronic respiratory diseases, n (%) | 68 (11.1%) |
| HIV/AIDS, n (%) | 1 (0.2%) |
| Severe chronic liver disease, n (%) | 5 (0.8%) |
| Rheumatological disease, n (%) | 4 (0.6%) |
| Dementia, n (%) | 3 (0.5%) |
| Malnutrition, n (%) | 0 |
| Neurological disorder, n (%) | 5 (0.8%) |
| Chronic hematological disease, n (%) | 3 (0.5%) |
| Immunodepression, n (%) | 6 (1%) |
| Current smoker | 65 (15.6%) |
| **Clinical signs before admission** |  |
| Fever, n (%) | 342 (55.6%) |
| Cough, n (%) | 473 (76.8%) |
| Dyspnea, n (%) | 251 (40.7%) |
| Rhinorrhea and/or nasal congestion | 7 (1.1%) |
| Chest pain or pressure | 19 (3.1%) |
| Myalgias | 76 (12.3%) |
| Arthralgia | 21 (3.4%) |
| Fatigue | 215 (34.9%) |
| Sore throat | 53 (8.6%) |
| Stomachache | 11 (1.8%) |
| Nausea/Vomiting | 40 (6.5%) |
| Diarrhea | 25 (4.1%) |
| Confusion, n (%) | 0 |
| Ageusia | 4 (0.6%) |
| Anosmia, n (%) | 3 (0.5%) |
| Systolic blood pressure median (IQR) | 120 / 20 |
| Diastolic blood pressure median (IQR) | 70 / 10 |
| Temperature median (IQR) | 36.6 / 0.8 |
| Respiration rate median (IQR) | 21 / 2.8 |
| Heart rate median (IQR) | 88 / 15 |
| Oxygen saturation median (IQR) | 94 / 4 |
| **Laboratory abnormalities at admission** |  |
| Leukocyte count/uL, median (IQR) | 5,950 / 3,050 |
| Neutrophil count/uL, median (IQR) | 3,820 / 2,590 |
| Lymphocyte count/uL, median (IQR) | 1,350 / 820 |
| Neutrophil/lymphocyte ratio | 2.87 / 2.41 |
| Platelet/lymphocyte ratio | 143.5 / 85.7 |
| Platelet count/uL, median (IQR) | 194,000 / 84,750 |
| Haemoglobin, g/dL, median (IQR) | 13 / 2 |
| Hematocrit, %, median (IQR) | 39 / 5.4 |
| Urea, mmol/L, median (IQR) | 28 / 13 |
| Creatinine, mg/dL, median (IQR) | 109 / 146 |
| Aspartate aminotransferase (AST), UI/L, median (IQR) | 31 / 19 |
| Alanine aminotransferase (ALT), UI/L, median (IQR) | 23 / 18 |
| Lactate dehydrogenase (LDH), UI/L, median (IQR) | 268 / 127 |
| Creatine phosphokinase (CPK), UI/L, median (IQR) | 99/146 |
| Albumin, g/L, median (IQR) | 37 / 5 |
| Ferritin, ng/L, median (IQR) | 155.5 / 243 |
| C-reactive protein (CRP), mg/L, median (IQR) | 42.8 / 74.5 |
| Troponin, ng/mL, median (IQR) | 4.1 / 5.5 |
| Procalcitonin, ng/mL, median (IQR) | 0.18 / 0.07 |
| D-dimer, mg/L, median (IQR) | 0.64 / 0.69 |
| **Chest CT** |  |
| Unilateral | 35 (6%) |
| Bilateral | 551 (94%) |
| **NEWS2 at D0** | Overall (n: ) |
| Median (min, max) | 4 (0-14) |
| Class I | 352 (57.6%) |
| Class II | 153 (25%) |
| Class III | 106 (17.3%) |
| **NEWS2 at D3** | Overall (n: ) |
| Median (min, max) | 3 (0-14) |
| Class I | 322 (72%) |
| Class II | 89 (19.9%) |
| Class III | 36 (8.1%) |
| **NEWS2 at D5** | Overall (n: ) |
| Median (min, max) | 3 (0-12) |
| Class I | 308 (75.7%) |
| Class II | 67 (16.5%) |
| Class III | 32 (7.9%) |
| **NEWS2 at D7** | Overall (n: ) |
| Median (min, max) | 3 (0-9) |
| Class I | 239 (78.9%) |
| Class II | 45 (14.9%) |
| Class III | 19 (6.3) |

Class I: 0-4 points
Class II: 5-6 points
Class III: 7-20 points

### 3.2 ICU admission or in-hospital mortality

The parameters associated with admission ICU and in-hospital death at D0, D3, D5, and D7 were NEWS2, lymphocyte count, neutrophil count, platelet count, NLR, PLR, CRP, procalcitonin, d-dimer, troponin, AST, urea, LDH, and albumin. The median and IQR values of the laboratory parameters and are available in Table-1.

**Table 1.** Median and interquartile range (IQR) values of the parameters

| Parameters | D0 |  | D3 |  | D5 |  | D7 |  |
| --- | --- | --- | --- | --- | --- | --- | --- | --- |
|  | Prognosis |  | Prognosis |  | Prognosis |  | Prognosis |  |
| **NEWS2** | **Poor** | **Good** | **Poor** | **Good** | **Poor** | **Good** | **Poor** | **Good** |
| IQR | 4 | 3 | 3 | 2 | 3 | 3 | 4 | 2 |
| Median | 6 | 4 | 5 | 3 | 6 | 3 | 7 | 3 |
| p value | 0.001 |  | 0.001 |  | 0.001 |  | 0.001 |  |
| **Leukocyte count** | **Poor** | **Good** | **Poor** | **Good** | **Poor** | **Good** | **Poor** | **Good** |
| IQR | 4020 | 2990 | 6745 | 2628 | 6383 | 2390 | 4223 | 2200 |
| Median | 6075 | 5930 | 7905 | 5800 | 7910 | 5880 | 7620 | 6220 |
| p value | 0.26 |  | 0.001 |  | 0.01 |  | 0.04 |  |
| **Lymphocyte count** | **Poor** | **Good** | **Poor** | **Good** | **Poor** | **Good** | **Poor** | **Good** |
| IQR | 538 | 810 | 508 | 750 | 295 | 793 | 786 | 1600 |
| Median | 1030 | 1390 | 805 | 1420 | 710 | 1505 | 685 | 1470 |
| p value | 0.001 |  | 0.001 |  | 0.001 |  | 0.001 |  |
| **Neutrophil count** | **Poor** | **Good** | **Poor** | **Good** | **Poor** | **Good** | **Poor** | **Good** |
| IQR | 4013 | 2410 | 5830 | 2038 | 6750 | 1910 | 3958 | 1810 |
| Median | 4645 | 3710 | 6565 | 3505 | 6520 | 3680 | 6475 | 3800 |
| p value | 0.001 |  | 0.001 |  | 0.001 |  | 0.001 |  |
| **Platelet count ×10<sup>3</sup>** | **Poor** | **Good** | **Poor** | **Good** | **Poor** | **Good** | **Poor** | **Good** |
| IQR | 83 | 82.75 | 132.5 | 120.75 | 73 | 163.5 | 74.5 | 160 |
| Median | 175 | 197 | 200.5 | 237 | 206.5 | 281 | 229.5 | 329 |
| p value | 0.04 |  | 0.01 |  | 0.001 |  | 0.01 |  |
| **NLR** | **Poor** | **Good** | **Poor** | **Good** | **Poor** | **Good** | **Poor** | **Good** |
| IQR | 4.54 | 2.11 | 8.28 | 1.84 | 7.36 | 1.86 | 10.65 | 1.64 |
| Median | 4.83 | 2.7 | 6.6 | 2.35 | 8.5 | 2.29 | 7.77 | 2.6 |
| p value | 0.001 |  | 0.001 |  | 0.001 |  | 0.001 |  |
| **PLR** | **Poor** | **Good** | **Poor** | **Good** | **Poor** | **Good** | **Poor** | **Good** |
| IQR | 129.8 | 78.2 | 173.1 | 101.6 | 154.9 | 119.8 | 280.8 | 135.2 |
| Median | 180.3 | 141.5 | 250.8 | 162 | 278.3 | 180.2 | 341.3 | 223.3 |
| p value | 0.001 |  | 0.001 |  | 0.001 |  | 0.01 |  |
| **CRP** | **Poor** | **Good** | **Poor** | **Good** | **Poor** | **Good** | **Poor** | **Good** |
| IQR | 99.8 | 65 | 139 | 60 | 115.3 | 50.3 | 139.6 | 48.6 |
| Median | 110.5 | 33.1 | 127 | 29.8 | 183 | 24 | 142.5 | 19 |
| p value | 0.001 |  | 0.001 |  | 0.001 |  | 0.001 |  |
| **Procalcitonin** | **Poor** | **Good** | **Poor** | **Good** | **Poor** | **Good** | **Poor** | **Good** |
| IQR | 0.64 | 0.05 | 0.79 | 0.05 | 1.08 | 0.03 | 0.88 | 0.05 |
| Median | 0.19 | 0.04 | 0.45 | 0.04 | 0.58 | 0.04 | 0.20 | 0.05 |
| p value | 0.001 |  | 0.001 |  | 0.001 |  | 0.01 |  |
| **D-dimer** | **Poor** | **Good** | **Poor** | **Good** | **Poor** | **Good** | **Poor** | **Good** |
| IQR | 1.06 | 0.64 | 3.82 | 0.76 | 5.97 | 0.95 | 4.47 | 1.11 |
| Median | 0.8 | 0.63 | 1.4 | 0.8 | 1.4 | 1 | 2.7 | 1.3 |
| p value | 0.03 |  | 0.001 |  | 0.01 |  | 0.04 |  |
| **Ferritin** | **Poor** | **Good** | **Poor** | **Good** | **Poor** | **Good** | **Poor** | **Good** |
| IQR | 416 | 218 | 840 | 174 | 4008 | 241 | 1631 | 190 |
| Median | 234 | 146 | 583 | 171 | 500 | 203 | 499 | 208 |
| p value | 0.01 |  | 0.001 |  | 0.01 |  | 0.16 |  |
| **Troponin** | **Poor** | **Good** | **Poor** | **Good** | **Poor** | **Good** | **Poor** | **Good** |
| IQR | 39 | 4.40 | 29.80 | 3.40 | 35.85 | 2.92 | 44.90 | 4.00 |
| Median | 14.6 | 3.9 | 12.5 | 3.2 | 27.5 | 3.3 | 19.1 | 4 |
| p value | 0.001 |  | 0.001 |  | 0.01 |  | 0.01 |  |
| **AST** | **Poor** | **Good** | **Poor** | **Good** | **Poor** | **Good** | **Poor** | **Good** |
| Mean | 23 | 19 | 41 | 24 | 24 | 21 | 27 | 22 |
| Median | 36.5 | 30 | 51 | 31 | 48 | 36 | 46.5 | 36 |
| p value | 0.001 |  | 0.001 |  | 0.001 |  | 0.04 |  |
| **ALT** | **Poor** | **Good** | **Poor** | **Good** | **Poor** | **Good** | **Poor** | **Good** |
| IQR | 13 | 19 | 34 | 26 | 28 | 32 | 30 | 37 |
| Median | 22.5 | 23 | 27 | 26 | 30.5 | 32 | 28.5 | 35.5 |
| p value | 0.86 |  | 0.13 |  | 0.95 |  | 0.31 |  |
| **Creatinine** | **Poor** | **Good** | **Poor** | **Good** | **Poor** | **Good** | **Poor** | **Good** |
| IQR | 0.52 | 0.30 | 0.80 | 0.25 | 1.14 | 0.25 | 0.97 | 0.20 |
| Median | 0.98 | 0.72 | 0.86 | 0.7 | 0.9 | 0.68 | 0.9 | 0.7 |
| p value | 0.001 |  | 0.001 |  | 0.02 |  | 0.09 |  |
| **Urea** | **Poor** | **Good** | **Poor** | **Good** | **Poor** | **Good** | **Poor** | **Good** |
| IQR | 30 | 12 | 34 | 12 | 46 | 12 | 51 | 13 |
| Median | 39.4 | 27 | 43.7 | 25 | 43.5 | 24 | 33 | 25.8 |
| p value | 0.001 |  | 0.001 |  | 0.001 |  | 0.01 |  |
| **LDH** | **Poor** | **Good** | **Poor** | **Good** | **Poor** | **Good** | **Poor** | **Good** |
| IQR | 197 | 114 | 250 | 137 | 151 | 128 | 196 | 138 |
| Median | 336 | 262 | 468 | 272 | 513 | 281 | 485 | 289 |
| p value | 0.001 |  | 0.001 |  | 0.001 |  | 0.001 |  |
| **CPK** | **Poor** | **Good** | **Poor** | **Good** | **Poor** | **Good** | **Poor** | **Good** |
| IQR | 210 | 124 | 360 | 80 | 263 | 58 | 457 | 55 |
| Median | 170 | 102 | 181 | 72 | 124.5 | 63 | 161.5 | 57 |
| p value | 0.02 |  | 0.001 |  | 0.08 |  | 0.01 |  |
| **Albumin** | **Poor** | **Good** | **Poor** | **Good** | **Poor** | **Good** | **Poor** | **Good** |
| IQR | 8 | 5 | 6 | 5 | 7 | 5 | 9 | 4 |
| Median | 33 | 37 | 31 | 36.5 | 27.5 | 35 | 30 | 34 |
| p value | 0.001 |  | 0.001 |  | 0.001 |  | 0.001 |  |
| **Haemoglobin** | **Poor** | **Good** | **Poor** | **Good** | **Poor** | **Good** | **Poor** | **Good** |
| IQR | 2.4 | 2.0 | 2.5 | 2.2 | 2.0 | 1.9 | 2.7 | 2.0 |
| Median | 13 | 13 | 11.95 | 12.6 | 11.25 | 12.4 | 11.7 | 12.5 |
| p value | 0.43 |  | 0.01 |  | 0.001 |  | 0.09 |  |
| **Hematocrit** | **Poor** | **Good** | **Poor** | **Good** | **Poor** | **Good** | **Poor** | **Good** |
| IQR | 7.1 | 5.1 | 5.7 | 5.2 | 4.8 | 6.0 | 6.1 | 5.8 |
| Median | 38.9 | 39 | 36.05 | 37.7 | 34.3 | 37.1 | 35.6 | 36.7 |
| p value | 0.24 |  | 0.01 |  | 0.001 |  | 0.16 |  |

(D0: Admission, D3: Day-3, D5: Day-5, D7: Day-7, IQR: Interquartile Range, NLR: Neutrophil/lymphocyte ratio, PLR: Platelet/lymphocyte ratio, AST: Aspartate aminotransferase, ALT: Alanine aminotransferase, LDH; Lactate dehydrogenase, CPK: Creatine phosphokinase, CRP: C-reactive protein)

### 3.3 Univariate analysis

In univariate analysis, among parameters associated with ICU admission or in-hospital death at D0, D3, D5, and D7, best predictors were NEWS2, procalcitonin, NLR, and albumin. Additionally, d-dimer (at D0, D3, and D7) and haemoglobin (at D3 and D5) were valuable predictors in univariate analysis (Table-2).

**Table 2.**
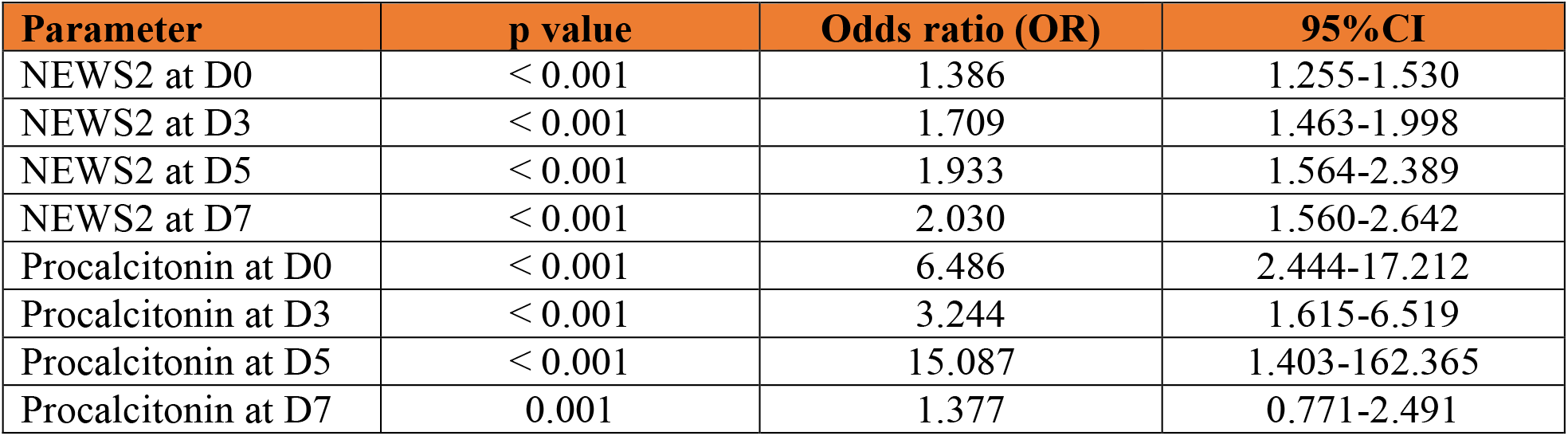
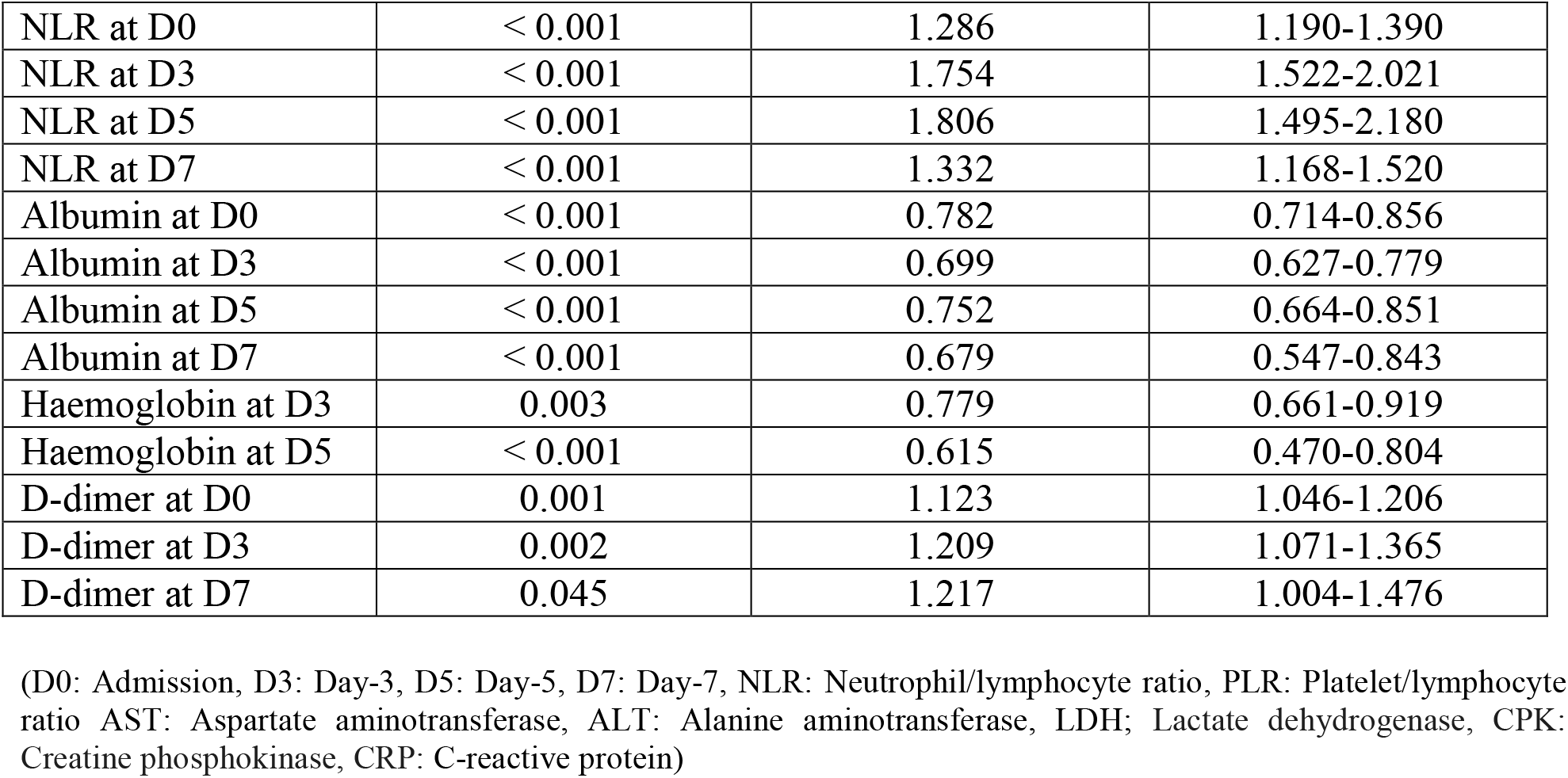
Univariate analysis of parameters associated with ICU admission and in-hospital death

| **Parameter** | **p value** | **Odds ratio (OR)** | **95%CI** |
| --- | --- | --- | --- |
| NEWS2 at D0 | < 0.001 | 1.386 | 1.255-1.530 |
| NEWS2 at D3 | < 0.001 | 1.709 | 1.463-1.998 |
| NEWS2 at D5 | < 0.001 | 1.933 | 1.564-2.389 |
| NEWS2 at D7 | < 0.001 | 2.030 | 1.560-2.642 |
| Procalcitonin at D0 | < 0.001 | 6.486 | 2.444-17.212 |
| Procalcitonin at D3 | < 0.001 | 3.244 | 1.615-6.519 |
| Procalcitonin at D5 | < 0.001 | 15.087 | 1.403-162.365 |
| Procalcitonin at D7 | 0.001 | 1.377 | 0.771-2.491 |
| NLR at D0 | < 0.001 | 1.286 | 1.190-1.390 |
| NLR at D3 | < 0.001 | 1.754 | 1.522-2.021 |
| NLR at D5 | < 0.001 | 1.806 | 1.495-2.180 |
| NLR at D7 | < 0.001 | 1.332 | 1.168-1.520 |
| Albumin at D0 | < 0.001 | 0.782 | 0.714-0.856 |
| Albumin at D3 | < 0.001 | 0.699 | 0.627-0.779 |
| Albumin at D5 | < 0.001 | 0.752 | 0.664-0.851 |
| Albumin at D7 | < 0.001 | 0.679 | 0.547-0.843 |
| Haemoglobin at D3 | 0.003 | 0.779 | 0.661-0.919 |
| Haemoglobin at D5 | < 0.001 | 0.615 | 0.470-0.804 |
| D-dimer at D0 | 0.001 | 1.123 | 1.046-1.206 |
| D-dimer at D3 | 0.002 | 1.209 | 1.071-1.365 |
| D-dimer at D7 | 0.045 | 1.217 | 1.004-1.476 |

(D0: Admission, D3: Day-3, D5: Day-5, D7: Day-7, NLR: Neutrophil/lymphocyte ratio, PLR: Platelet/lymphocyte ratio AST: Aspartate aminotransferase, ALT: Alanine aminotransferase, LDH; Lactate dehydrogenase, CPK: Creatine phosphokinase, CRP: C-reactive protein)

### 3.4 Correlation

Laboratory parameters correlated with NEWS2 at D0, D3, D5, and D7 were lymphocyte count, neutrophil count, NLR, PLR, CRP, procalcitonin, ferritin and urea (Suppl. Fig. 1. and Suppl. Fig. 2.)

**Supp. Figure 1.**
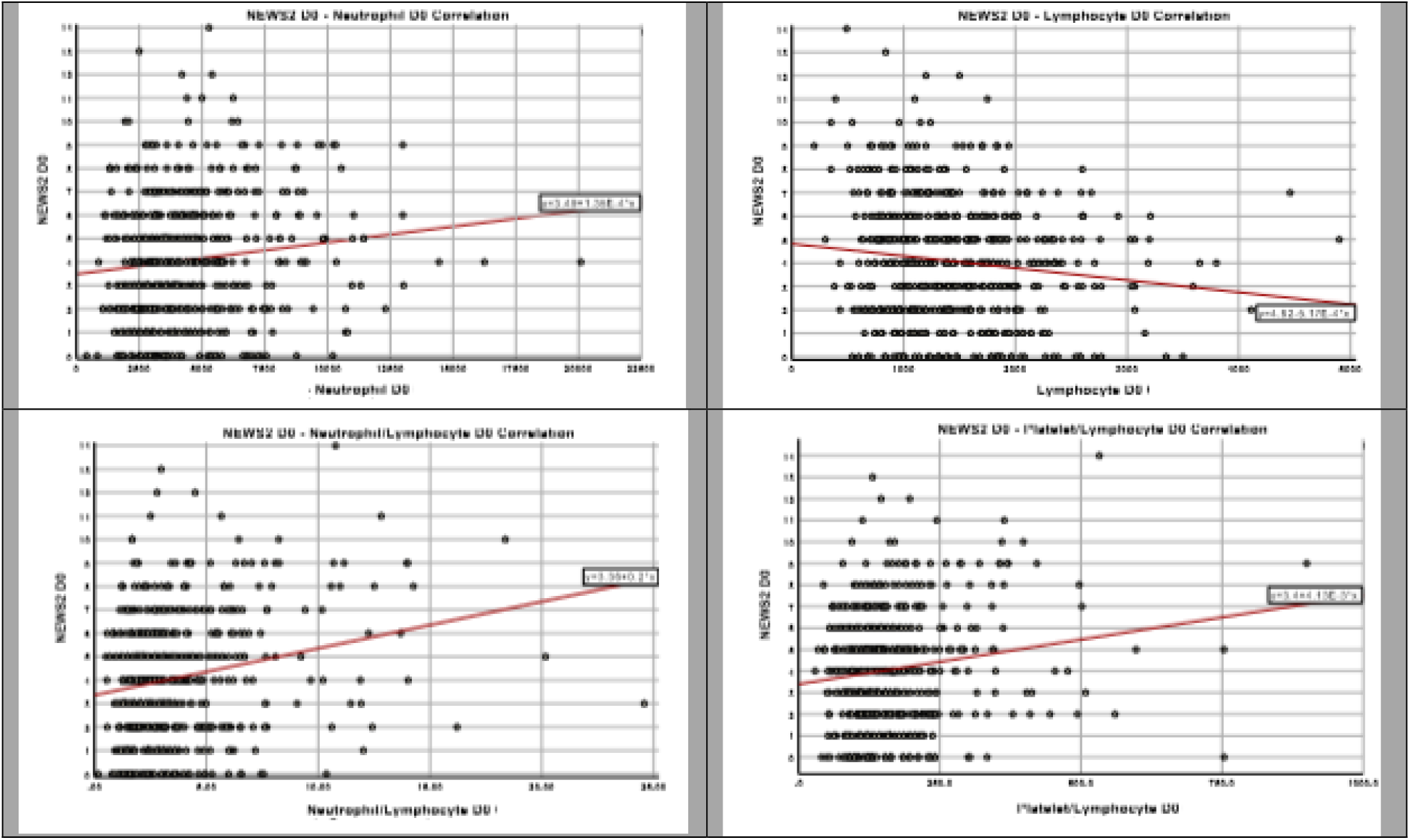
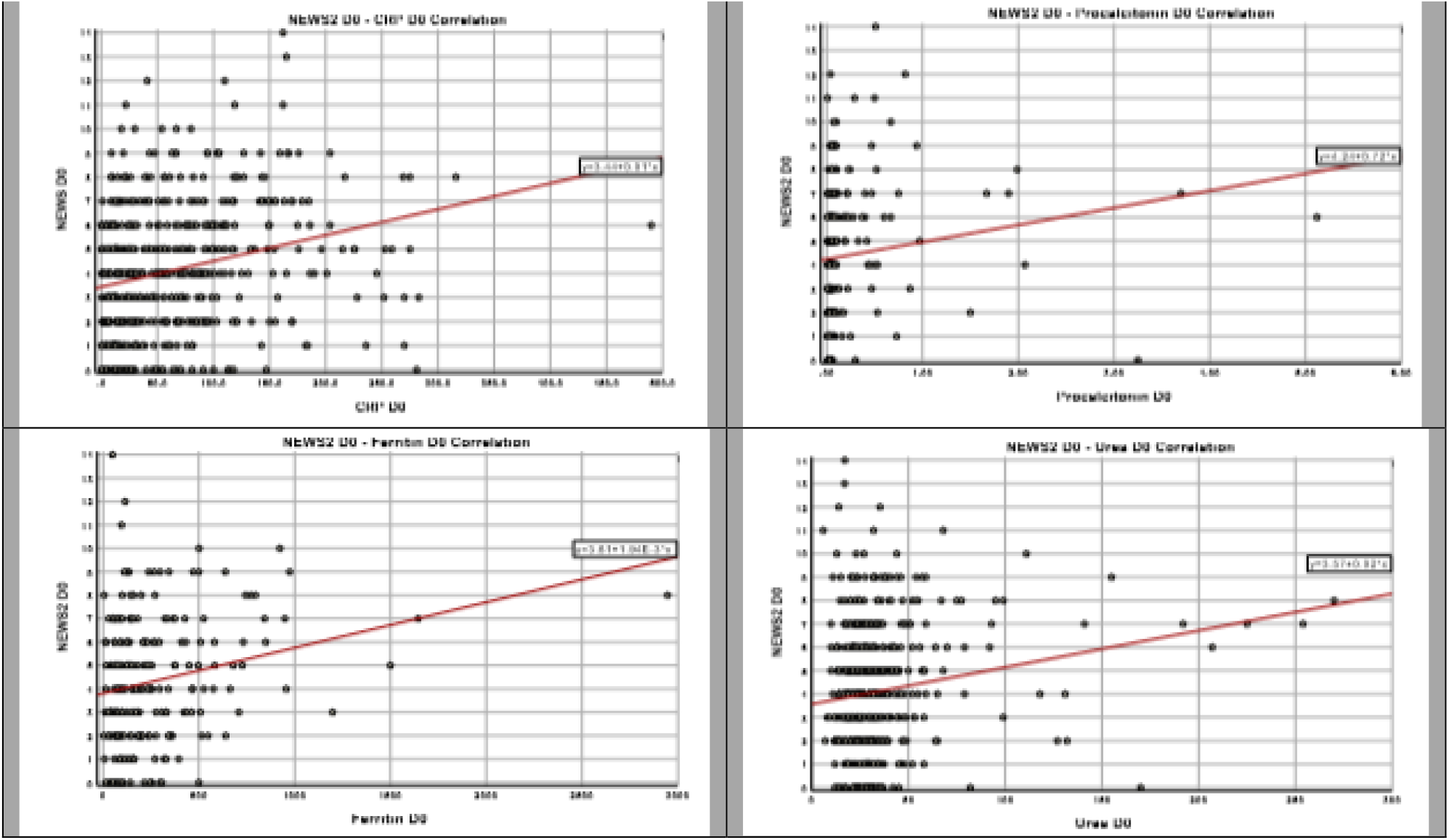
Correlation between NEWS2 and lymphocyte count, neutrophil count, NLR, PLR, CRP, procalcitonin, ferritin and urea at D0

**Supp Figure 2.**
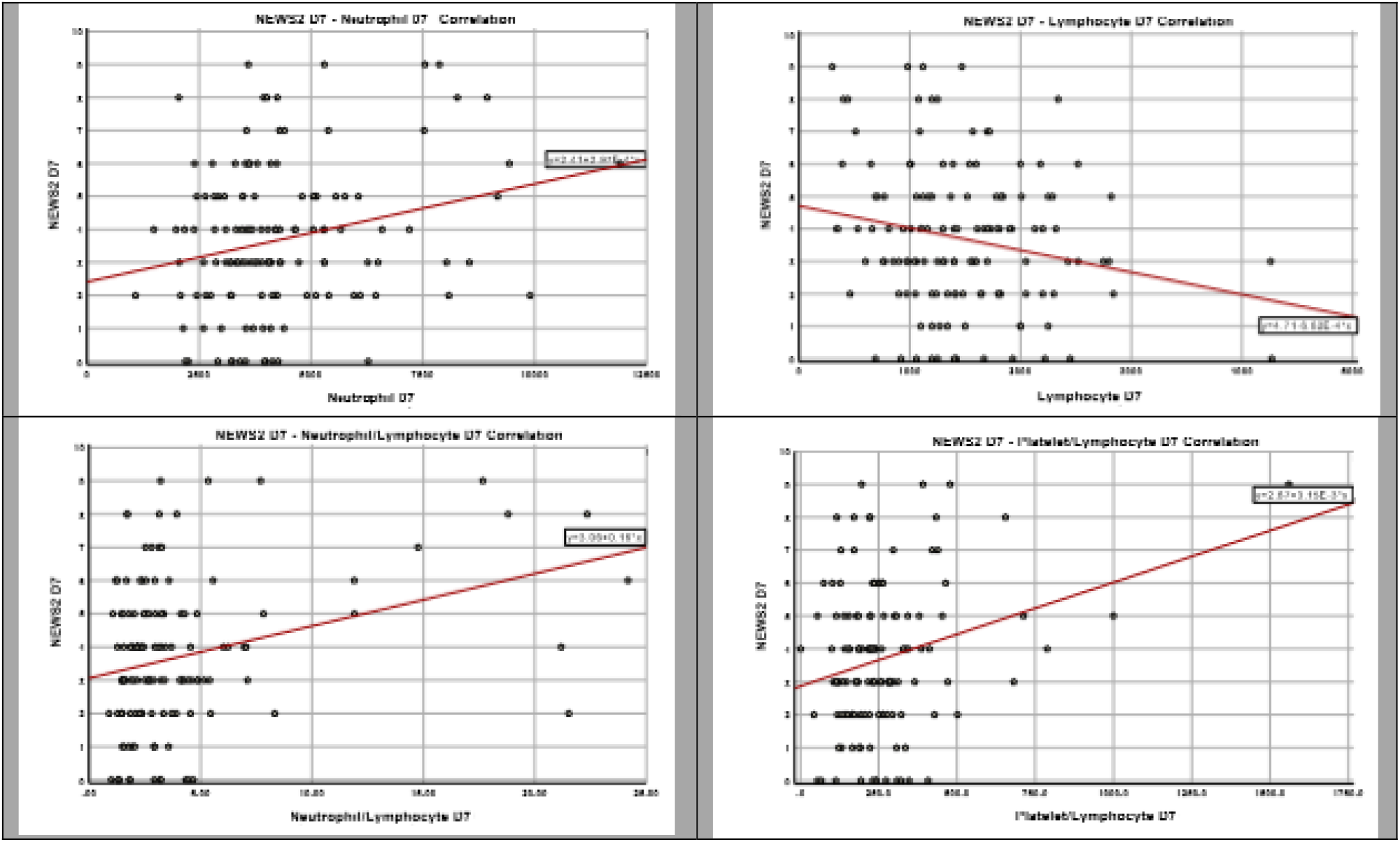
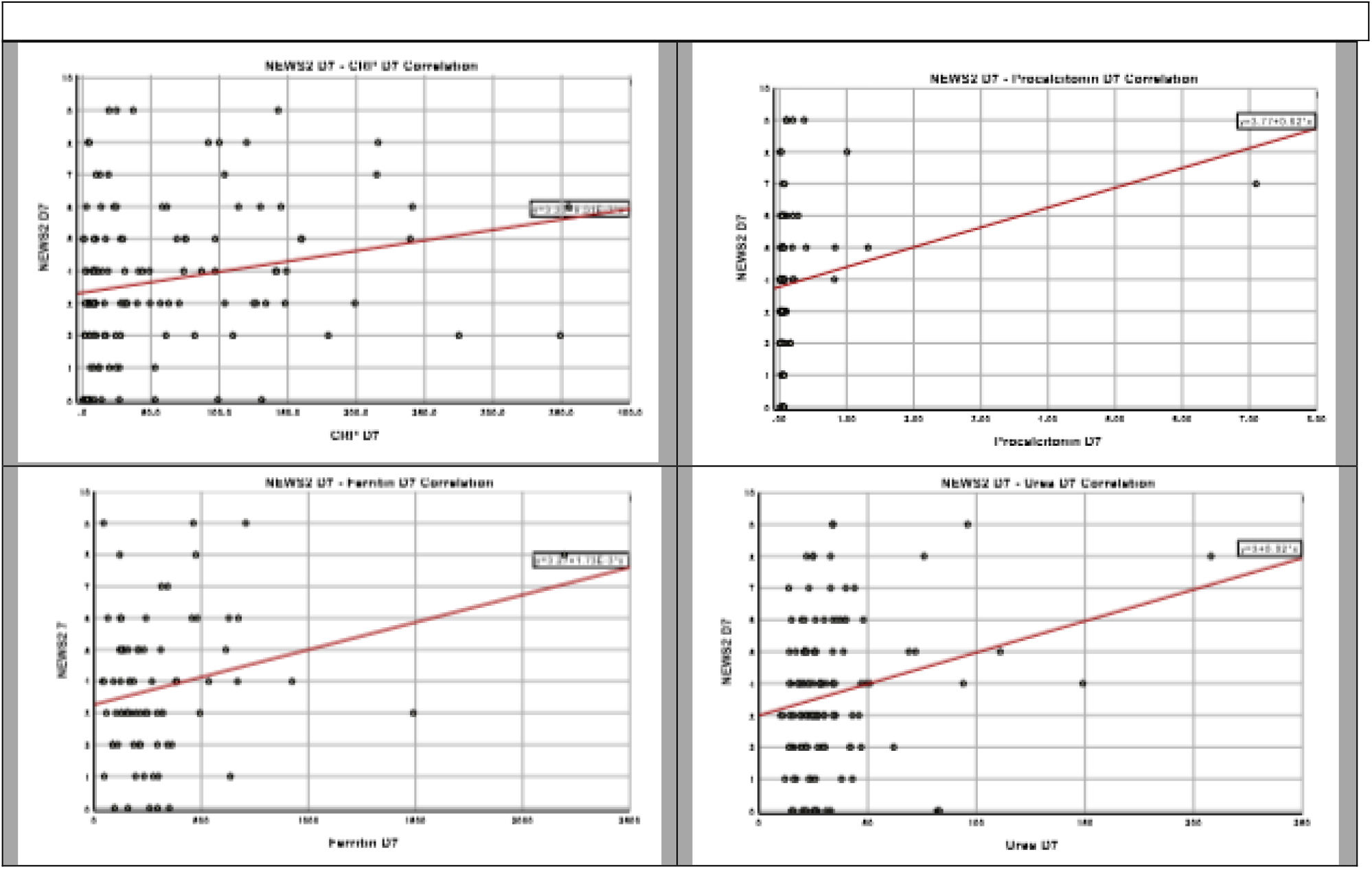
Correlation between NEWS2 and lymphocyte count, neutrophil count, NLR, PLR, CRP, procalcitonin, ferritin and urea at D7

### 3.5 ROC Curves

ROC curves of NEWS2 at D0, D3, D5 and D7 to predict clinical deterioration are shown in Fig. 1. At D0, AUC curve was 0.726 (95% CI 0.669-0.784), with the best cut off of 4.5, sensitivity of 71.1% and specificity of 61.7% (p<0.001). At D3, AUC curve was 0.798 (95% CI 0.740-0.856), with the best cut-off of 4.5, sensitivity of %69.6, and specificity of %78 (p<0.001). At D5, AUC curve was 0.833 (95% CI 0.755-0.911), with the best cut-off of 4.5, sensitivity of 72.4% and specificity of 79.4% (p<0.001). At D7, AUC curve was 0.842 (95% CI 0.739-0.945), with the best cut-off of 3.5, sensitivity of 88.2% and specificity of 64.7% (p<0.001).

ROC curves of procalcitonin at D0, D3, D5 and D7 to predict clinical deterioration are shown in Fig. 2. At D0, AUC curve was 0.824 (95% CI 0.755-0.893), with the best cut-off of 0.065, sensitivity of 81.1% and specificity of 67.9% (p<0.001). At D3, AUC curve was 0.896 (95% CI 0.837-0.956), with the best cut-off of 0.125, sensitivity of 80.6% and specificity of 89.3% (p<0.001). At D5, AUC curve was 0.967 (95% CI 0.928-1.000), with the best cut-off of 0.155, sensitivity of 87.5% and specificity of 96.7% (p<0.001). At D7, AUC curve was 0.823 (95% CI 0.609-1.000), with the best cut-off of 0.120, sensitivity of 85.7 % and specificity of 88.2% (p<0.004). ROC curves of albumin at D0, D3, D5 and D7 to predict clinical deterioration are shown in Fig 3. At D0, AUC curve was 0.746 (95% CI 0.656-0.835), with the best cut-off of 35.5, sensitivity of 69.2% and specificity of 66.7% (p<0.001). At D3, AUC curve was 0.868 (95% CI 0.816-0.919), with the best cut-off of 33.5, sensitivity of 79.8% and specificity of 78.3% (p <0.001). At D5, AUC curve was 0.887 (95% CI 0.814-0.960), with the best cut-off of 31.5, sensitivity of 89.3% and specificity of 70% (p<0.001). At D7, AUC curve was 0.896 (95% CI 0.813-0.979), with the best cut-off of 32.5, sensitivity of 78.9% and specificity of 88.9% (p<0.001).

ROC curves of neutrophil/lymphocyte ratio at D0, D3, D5 and D7 to predict clinical deterioration are shown in Fig. 4. At D0, AUC curve was 0.752 (95% CI 0.694-0.810), with the best cut-off of 40.95, sensitivity of 64.5% and specificity of 77% (p<0.001). At D3, AUC curve was 0.893 (95% CI 0.843-0.942), with the best cut-off of 38.60, sensitivity of 86.7% and specificity of 81.3% (p<0.001). At D5, AUC curve was 0.939 (95% CI 0.899-0.979), with the best cut-off of 58.6, sensitivity of 79.2% and specificity of 94.8% (p<0.001). At D7, AUC curve was 0.911 (95% CI 0.834-0.988), with the best cut-off of 58.45, sensitivity of 81.3% and specificity of 95% (p<0.001).

**Figure 1.**
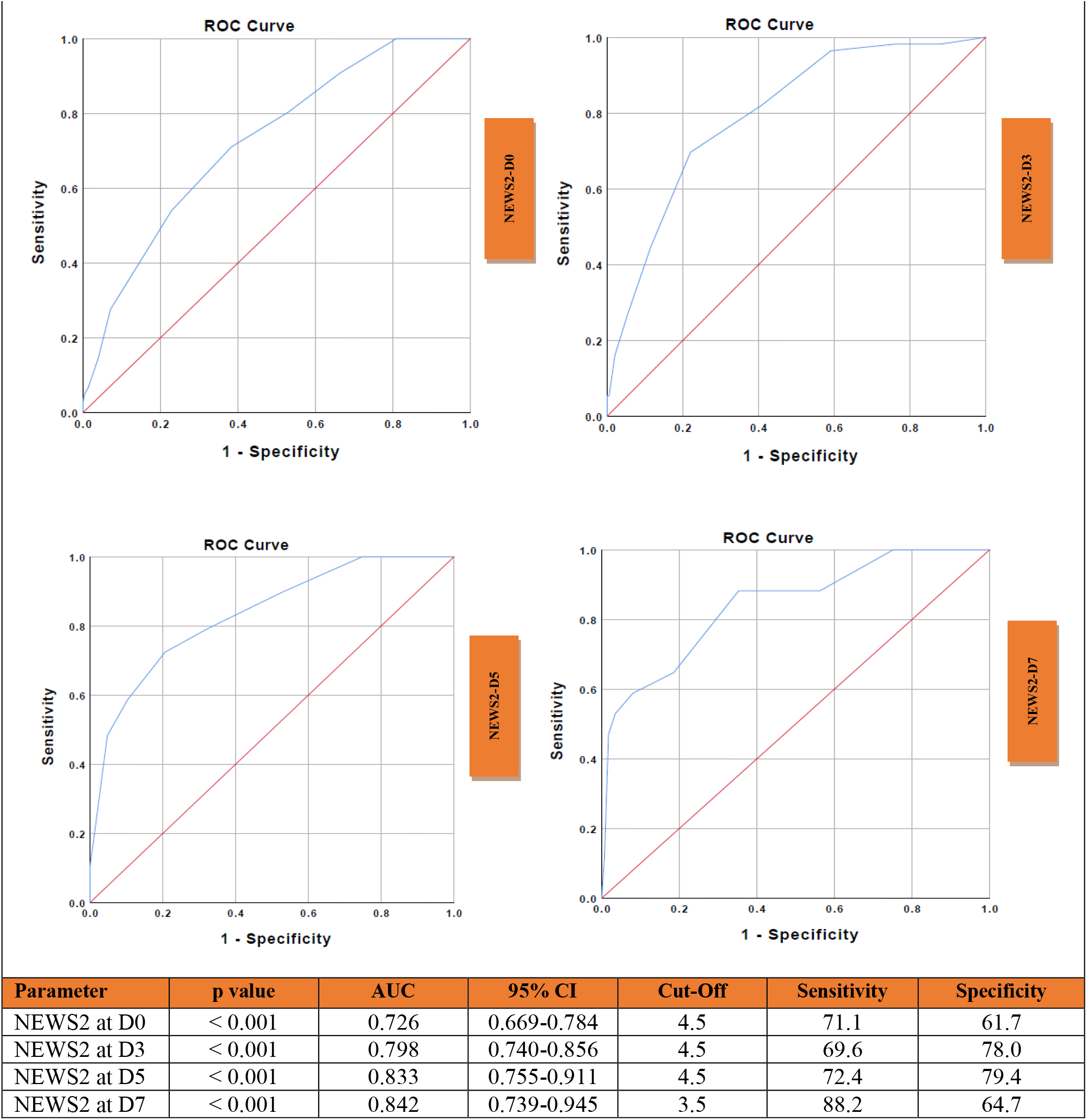
Receiver Operating Characteristic (ROC) curve and performance value for the best cut off for: **A**: NEWS2 at admission using clinical deterioration **B**: NEWS2 at D3 using clinical deterioration **C**: NEWS2 at D5 using clinical deterioration **D**: NEWS2 at D7 using clinical deterioration

**Figure 2.**
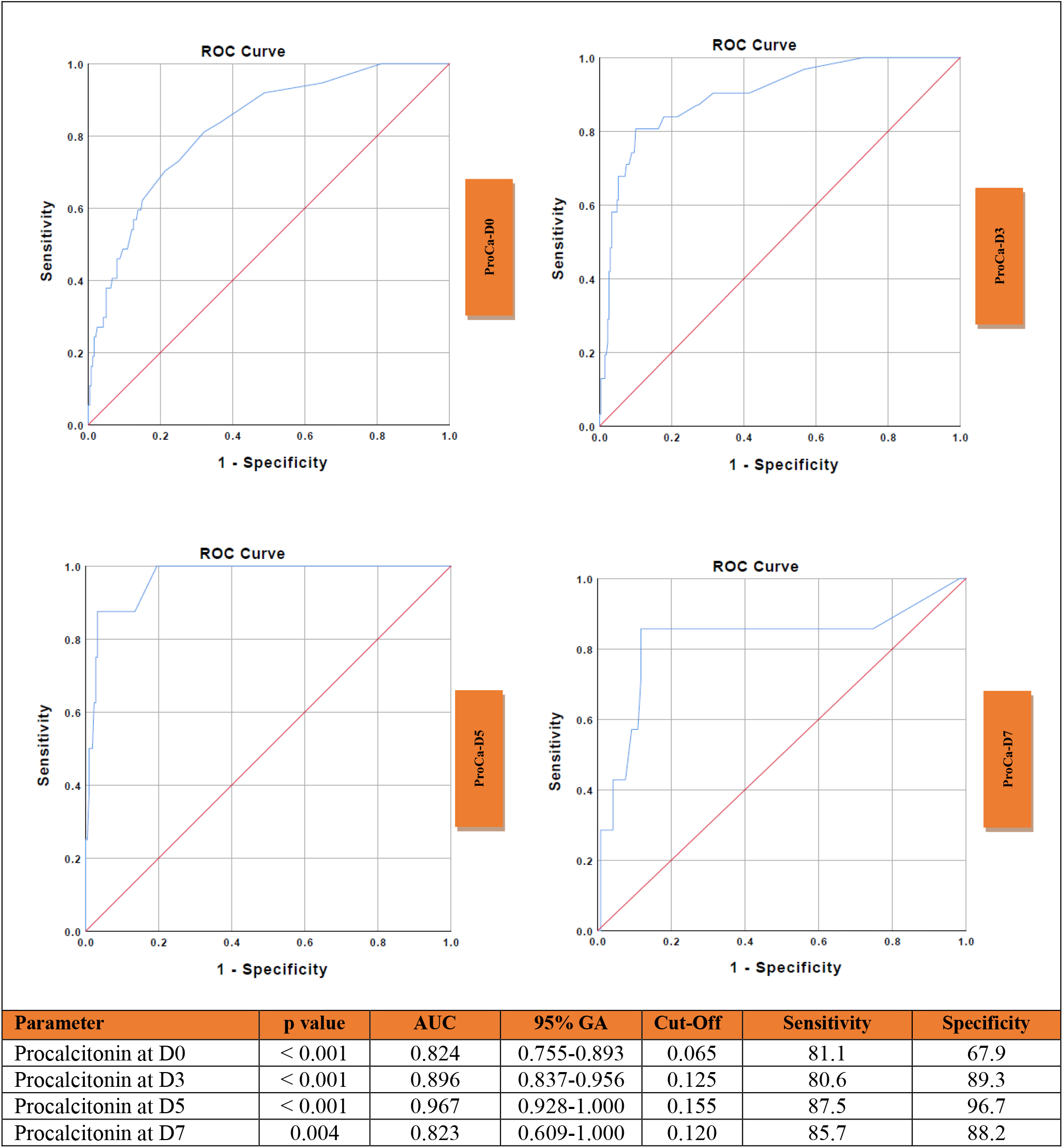
Receiver Operating Characteristic (ROC) curve and performance value for the best cut off for: **A**: procalcitonin at admission using clinical deterioration. **B**: procalcitonin at D3 using clinical deterioration. **C**: procalcitonin at D5 using clinical deterioration. **D**: procalcitonin at D7 using clinical deterioration.

**Figure 3.**
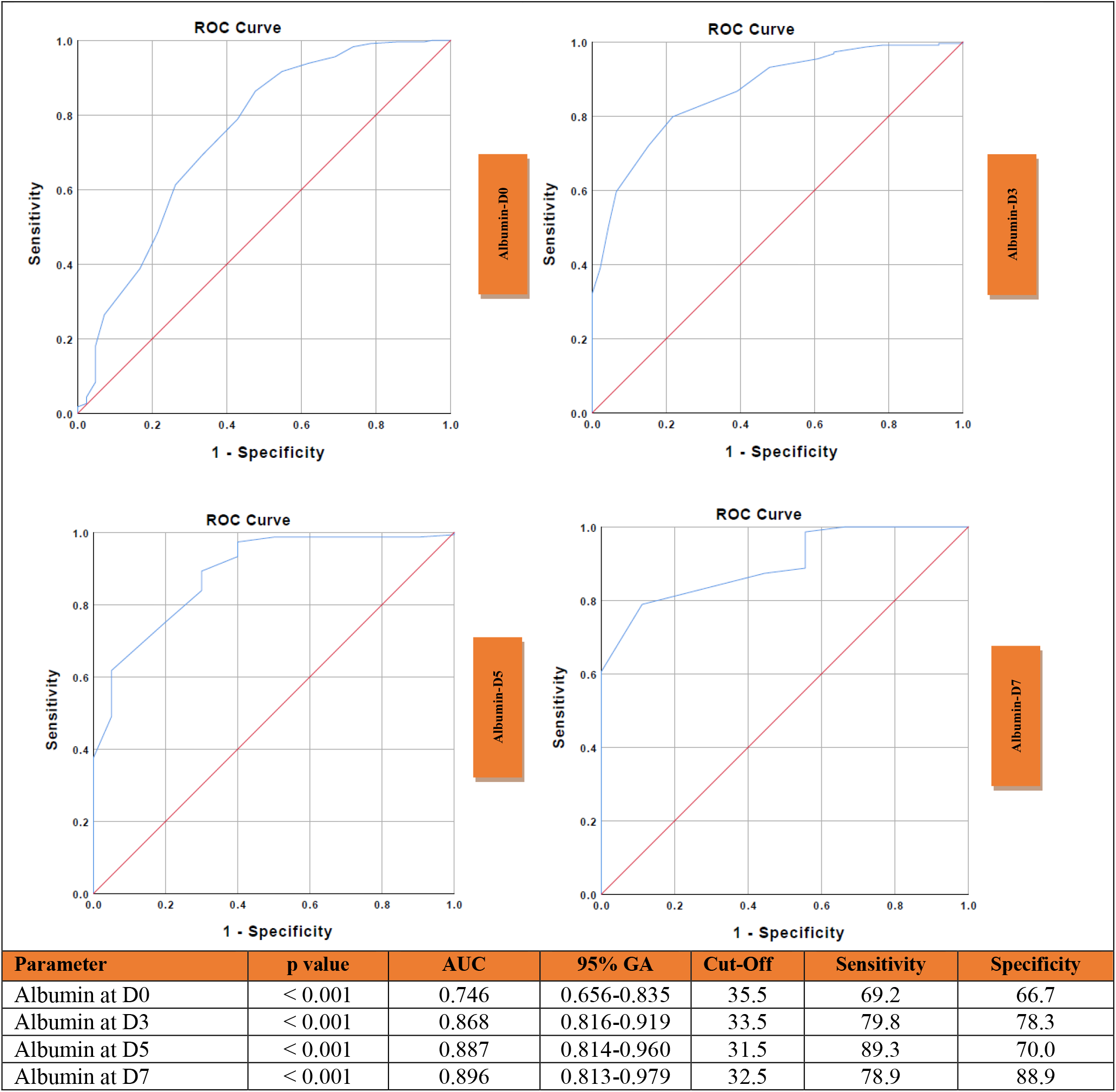
Receiver Operating Characteristic (ROC) curve and performance value for the best cut off for: **A:** albumin at admission using clinical deterioration **B:** albumin at D3 using clinical deterioration **C:** albumin at D5 using clinical deterioration **D:** albumin at D7 using clinical deterioration.

**Figure 4.**
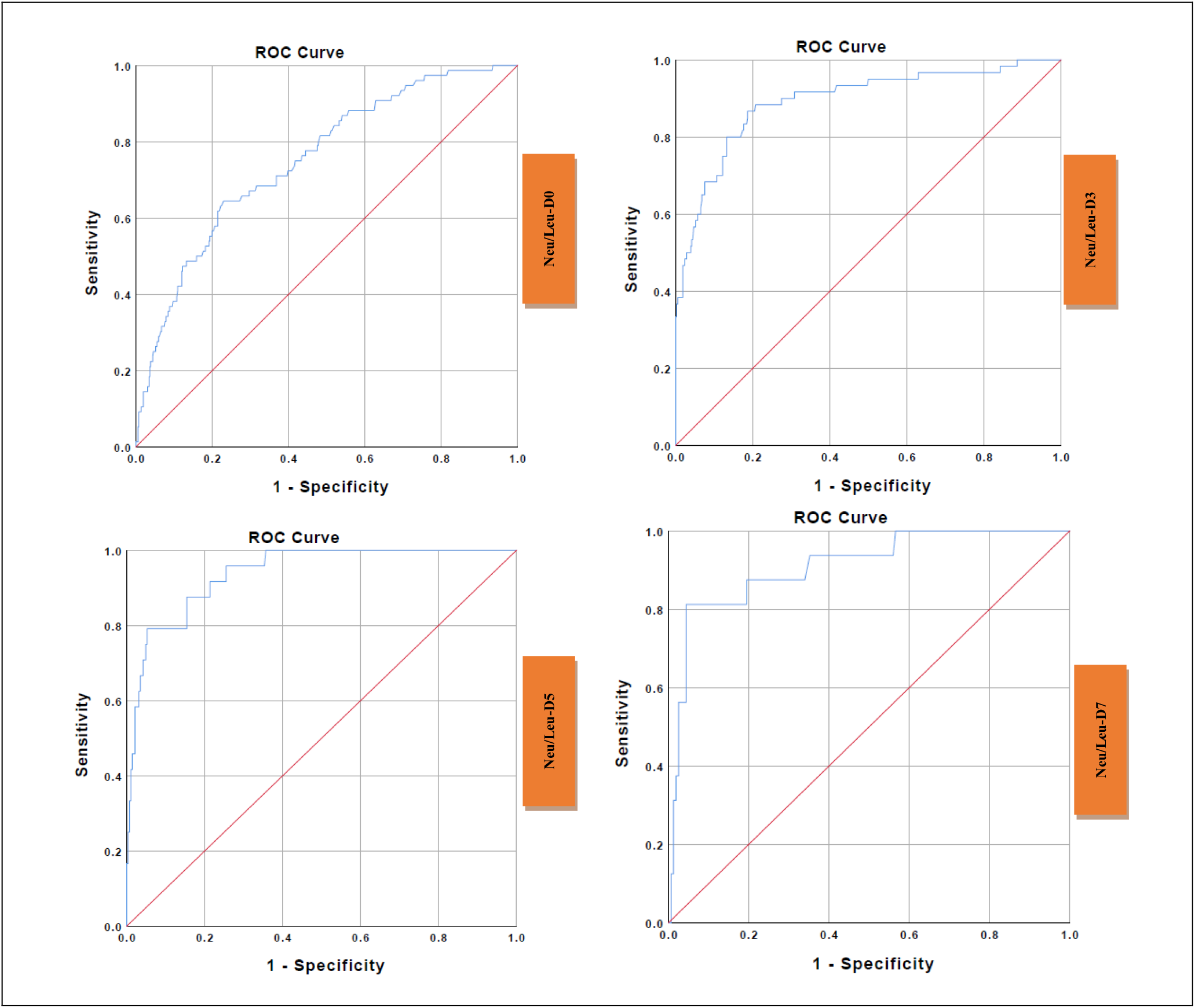
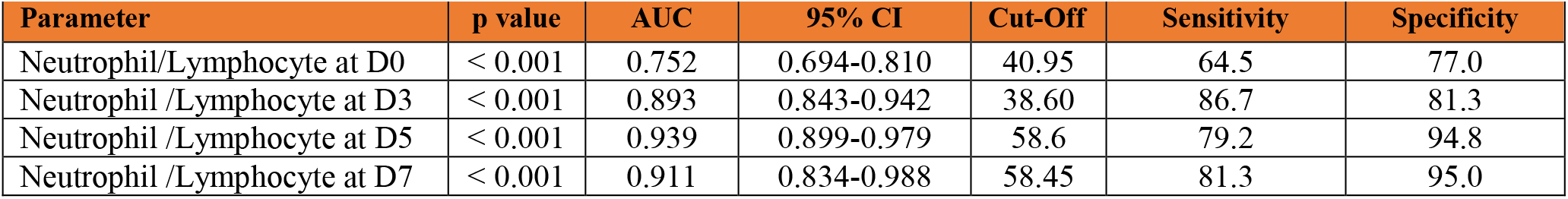
Receiver Operating Characteristic (ROC) curve and performance value for the best cut off for: **A**: neutrophil/lymphocyte ratio (NLR) at admission using clinical deterioration **B**: NLR at D3 using clinical deterioration **C**: NLR at D5 using clinical deterioration **D**: NLR at D7 using clinical deterioration

## 4. DISCUSSION

In this study, we presented a detailed analysis of the NEWS2 score and laboratory parameters in hospitalized patients with COVID-19. Our results showed that NEWS2, procalcitonin, NLR, and albumin were the best predictors for clinical deterioration (ICU admission or in-hospital death) at D0, D3, D5, and D7. Procalcitonin had the highest odds ratio for clinical deterioration at D0, D3, D5, and D7 in univariate analysis. ROC analyses showed that NEWS2 at D7, procalcitonin at D5, albumin at D7, and NLR at D5 had highest AUC values. Additionally, we detected a strong correlation between NEWS2 and laboratory parameters including lymphocyte count, neutrophil count, NLR, PLR, CRP, procalcitonin, ferritin, and urea at D0, D3, D5, and D7.

Early and accurate discrimination of need for ICU improves the clinical course of COVID-19 and reduce unnecessary use of ICU beds. To date limited data exists on the NEWS2 and laboratory parameters in patients with COVID-19. There are several published studies on the use of NEWS2 in COVID-19 patients (2-4, 7-12). However, most studies evaluate NEWS2 at admission only (10-13). In the study of Sze et al., they suggested that NEWS2 score was not a valuable tool to predict clinical deterioration in elderly patients with COVID-19 (14). However, they reported the results of only 17 elderly patients. Kim et al. showed that NEWS2 scores on D0 significantly differed in non-critical and critical patients (2.6 ± 2.6 vs. 8.2 ± 3.3, p<0.001) (9). In the study of Volff et al., the AUC value of NEWS score to predict ICU admisson or death was 0.74, in consistent with our result. However, the AUROC values of NEWS2 at D3, D5, and D7 were 0.798, 0.833, and 0.842 whereas NEWS2 at D0 was not accurate (AUROC<0.750) (7). Similarly, Sixt et al. showed that AUROC values of NEWS2 was 0.74 at D0, with a best cut-off of 6 and was 0.98 at D7, with a best cut-off of 7. They reported high sensitivity and specificity at D7 (%92 and %97, respectively) (8).

Due to the physiopathological changes, deterioration in different laboratory parameters occurs while the disease progresses. Therefore, laboratory parameters are commonly used for assessing disease severity. Lagadinou et al. found an association between COVID-19 severity and the following laboratory parameters NLR, LDH, d-dimers, CRP, fibrinogen, and ferritin (15). In the study of Xu et al., procalcitonin, CRP, and NLR were valuable predictors for COVID-19 mortality. They showed that the AUC from highest to lowest was combined effect> CRP > procalcitonin > NLR, respectively (16). Liao et al. reported that NLR, thrombocytopenia, prothrombin time, and D-dimer were associated with death. They showed that increased NLR (≥9·13) was associated with 5-fold increased mortality risk (17). Similarly, in our study, increased NLR was associated with OR = 1.286-fold at D0, OR = 1.754-fold at D3, OR = 1.806-fold at D5, and OR = 1.332-fold at D7 increased mortality risk in univariate analysis. In a meta-analysis, Elshazli et al. demonstrated that higher levels of leukocyte (OR = 5.21), neutrophil (OR = 6.25), D-dimer (OR = 4.19), and prolonged PT (OR = 2.18) was associated with ICU admission. IL-6 (OR = 13.87), CRP (OR = 7.09), D-dimer (OR = 6.36), and neutrophils (OR = 6.25) had the highest OR for mortality (18). In a meta-analysis, Lippi et al showed that increased procalcitonin values are associated with a nearly 5-fold higher risk for need for ICU or use of mechanical ventilation (OR, 4.76; 95% CI, 2.74–8.29) (19). Xu et al. found that procalcitonin (≥ 0.10 ng/mL, HR=12.82), CRP (≥ 52.14 mg/L, HR=12.30), and NLR (≥ 3.59, HR=8.6) had higher HRs of 12.82, 12.30 and 8.6 for mortality, respectively. Additionally, procalcitonin (≥ 0.10 ng/mL) and CRP (≥ 52.14 mg/L) but not NLR exhibited independent increasing risks of mortality, with HRs of 52.68 (95% CI: 1.77–1571.66) and 5.47 (95% CI: 1.04–28.72), respectively (16).

In the study of Shang et al., Spearman’s rank correlation analysis revealed that leukocyte, neutrophil, CRP, procalcitonin, and LDH were positively correlated and albumin was negatively correlated with mortality in patients with receiving maintenance hemodialysis. Additionally, they showed that CRP had the highest AUROC value (0.895), and the values of AUROC of neutrophil count, LDH, leukocyte, albumin, and procalcitonin were 0.813, 0.758, 0.757, 0.743, and 0.728, respectively (20). In contrast, we found that procalcitonin was the best predictor for clinical deterioration our study. The optimal cut-off value of procalcitonin at D0, D3, D5, and D7 were 0.065 ng/mL, 0.125 ng/mL, 0.155 ng/mL, and 0.120 ng/mL, and the sensitivity and specificity to predict clinical deterioration were 81.1% and 67.9% on D0, 80.6 % and 89.3 % on D3, 87.5 % and 96.7 % on D5, and 85.7 % and 88.2 % on D7 respectively.

Procalcitonin is not well studied for COVID-19 cases. However, some studies suggested that increased procalsitonin levels were found to be associated with the disease severity in patients with COVID-19. A meta-analysis showed that severe patients with COVID-19 had increased procalcitonin levels (18, 19). Similarly, we found that procalcitonin was the best prognostic parameter for the clinical deterioration in our study. Elevated procalcitonin levels could be associated with acute secondary bacterial pnemonia or systemic secondary bacterial infection in patients with COVID-19 due to the production and release into the circulation from procalcitonin producing extrathyroidal tissues. (21) In our study, despite elevated procalcitonin levels, this elevation was limited. In a previous study by Xu et al., they suggested that a limited increase in procalcitonin levels could be associated with increased interferon-gamma (16).

Low serum albumin in studies with COVID-19 patients levels are suggested to be associated with an increased risk of mortality (22-25) In consistent with other studies, our results confirm that albumin is a valuable predictor for ICU admission or in-hospital death. Albumin is a negative acute phase reactant produced in the liver and causes down-regulation of the expression of ACE-2 receptors which play a role in the cell entry mechanism of SARS-CoV-2. Liu et al. reported that albumin was associated with clinical deteroration and significantly higher in patients with the improvement/stabilization than in those with disease progression (36.62 ± 6.60 vs. 41.27 ± 4.55 g/L, p=0.006) (24). In the study of Aziz et al., mean albumin at D0 was 3.50 g/dL (CI 3.26–3.74 g/dL) in severe group and 4.05 g/dL (CI 3.82–4.27 g/dL) in non-severe group (p < 0.001). They reported that hypoalbuminemia was associated with 12.6-fold increased risk of mortality (25).

An increase in neutrophils and a decrease in lymphocytes have been found in various studies. Some studies have shown that NLR may be an important indicator for the severity of COVID -19 patients. Yan et al. showed that NLR was significantly correlated with all-cause in-hospital mortality (OR 44.351; 95% CI 4.627-425.088) (26). The NLR reflects the balance between the innate and adaptive immune systems (30) and increased NLR levels were found to be associated with clinical deterioration in COVID-19 (16).

This study has also several limitations. First, it was retrospectively conducted in a single-center. Second, this study had a small sample size and a control group was not included. The generalizability of our results may be limited. Thus, we need new large scale studies providing important information to better understand COVID-19 pandemic. Our study has also several strengths. First, we were able to admit all critically ill patients requiring intensive care to the ICU during the first months of pandemic. This prevent a selection bias. Second, longitudinally evaluation of the association between clinical deterioration and the dynamic changes of laboratory parameters was performed, since we regularly monitored laboratory parameters during the clinical course.

## Conclusion

This study provides a list of several laboratory parameters correlated with NEWS2 and potential predictors for ICU admission or in-hospital death during the clinical course of COVID-19. NEWS2, procalcitonin, NLR, and albumin have a high accuracy to predict clinical outcomes/disease progression in hospitalized patients and should be considered in the clinical decision of ICU admission. In conclusion, dynamic monitoring of NEWS2 and laboratory parameters is vital for improving clinical outcomes.

## Data Availability

The data that support the findings of this study are available from the corresponding author upon reasonable request.

## Acknowledgements

The authors acknowledge all healthcare professionals who contribute to the care of our patients.

## Author Contributions

GT proposed the concept, designed the study, wrote the protocol, and managed the study. GT, SS, IYN, and MY, performed the statistics, interpreted the data, and wrote the manuscript. GT, SS, HKK, AB, AKC, BC, EZ, GT, HA, IYN, MY, YK were involved in collecting the data. MMS, SSY, FP, GS performed a critical review of the manuscript. All authors provided inputs for revision of the manuscript. SS communicated with the journal and addressed comments from reviewers. All authors contributed to data acquisition, data analysis, or data interpretation, and reviewed and approved the final version.

## Ethics Statement

All procedures performed in studies involving human participants were in accordance with the ethical standards of the Ethics Committee of Haseki Training and Research Hospital and national research committee and with the ethical standards of the Declaration of Helsinki. Written informed consent was waived, given the retrospective nature of this study.

